# Transmission dynamics of SARS-COV-2 in China: impact of public health interventions

**DOI:** 10.1101/2020.03.24.20036285

**Authors:** Wenbao Wang, Yiqin Chen, Qi Wang, Ping Cai, Ye He, Shanwen Hu, Yan Wu, Zuxiong Huang, Wenxiang Wang

## Abstract

COVID-19 has become a global pandemic. However, the impact of the public health interventions in China needs to be evaluated. We established a SEIRD model to simulate the transmission trend of China. In addition, the reduction of the reproductive number was estimated under the current forty public health interventions policies. Furthermore, the infection curve, daily transmission replication curve, and the trend of cumulative confirmed cases were used to evaluate the effects of the public health interventions. Our results showed that the SEIRD curve model we established had a good fit and the basic reproductive number is 3.38 (95% CI, 3.25–3.48). The SEIRD curve show a small difference between the simulated number of cases and the actual number; the correlation index (H^2^) is 0.934, and the reproductive number (R) has been reduced from 3.38 to 0.5 under the current forty public health interventions policies of China. The actual growth curve of new cases, the virus infection curve, and the daily transmission replication curve were significantly going down under the current public health interventions. Our results suggest that the current public health interventions of China are effective and should be maintained until COVID-19 is no longer considered a global threat.

## 1. Introduction

On Dec 8, 2019, in Wuhan, the capital city of Hubei Province, China, an atypical pneumonia (later named COVID-19 by the WHO) broke out. The pneumonia was discovered to be similar to the SARS outbreak of 2003. The clinical symptoms of the patients were fever, fatigue, dry cough, and gradual dyspnea. The severe cases suffered acute respiratory distress syndrome, septic shock, serious metabolic acidosis, and coagulation dysfunction[1-2]. On Dec 12, the Wuhan Municipal Health Commission (WMHC) reported 27 cases (seven of which were severe), and epidemiological studies showed that most of the patients had a history of exposure in the South China seafood market in Wuhan [3]. Subsequently, a novel coronavirus strain named SARS-COV-2 by the International Committee on Taxonomy of Viruses was isolated from these patients and its genome was sequenced [4]. The sequence similarity with the SARS coronavirus was 79.5% [5], and it was proposed that the intermediate hosts are wild animals such as snakes and bats [5-6]. At present, human-to-human transmission patterns such as family aggregation have emerged [7-8]. By Mar 5, 2020, the total number of confirmed cases in China has reached 63,420 (excluding clinically confirmed cases), and the total number of deaths has reached 3,042. The spread speed and scale of this epidemic disease far exceed those of SARS in 2003, which brought great harm and had a serious impact on the economy and society worldwide. At present, China is trying its best to treat COVID-19 and has controlled it well; however, until now, no vaccines or targeted drugs are available. Therefore, the most effective interventions are epidemic publicity and isolation, and preventing the spread of the disease is particularly important [9-10]. However, the effects of these public health interventions on the transmission dynamics of SARS-COV-2 of China are need to evaluated. In addition, only a few studies have been published to calculate the basic reproductive number, based on a few early cases [11-12]. However, the early high missed diagnosis and misdiagnosis rates may increase the error probability of model prediction. Therefore, it is important to further explore the transmission dynamics of COVID-19 with more recent and more complete data, which can provide more reliable support for the development and implementation of public health interventions.

As early as in 1911, Ross used a mathematical model to model the malaria epidemic in epidemiological studies [13]. In 1926, Kermack and McKendrick established the famous susceptible-infectious-recovered (SIR) model [14-15]. For the studies of SARS in 2003, many studies built models based on the SIR model [16-17]. With the improvement of SARS case reports and the known ecology of SARS disease, some studies built a transmission dynamics model called the susceptible-exposed-infectious-recovered (SEIR) four-compartment model [18], and even a susceptible-exposed-infectious-recovered-death (SEIRD) five-compartment model, and applied it to the transmission dynamics of other infectious diseases [19]. Therefore, the aim of this study was (1) to explore the transmission dynamics of SARS-COV-2 by using the SEIRD model and (2) to evaluate the impacts of the existing public health interventions, so as to provide a theoretical framework for the development and implementation of public health interventions in the world.

## 2. Methods

### 2.1 Data sources

Due to the low reliability of early diagnoses of COVID-19 caused by SARS-COV-2 and the unknown intermediate animal host, the epidemic scale of COVID-19 in the early period in Wuhan is not clear. Therefore, we used the incidence of COVID-19 caused by SARS-COV-2 confirmed by RT-PCR [20] in the population moving out from Wuhan to eight other provinces in China to reverse estimate the epidemic scale in Wuhan. A similar method was used in a previous study [11]. According to the current epidemic scale (Figure 1) and the migration data from the Baidu migration map, the data of the eight provinces to which the highest number of people moved from Wuhan during the period from Jan 14 to Jan 23 were collected. The incidence of COVID-19 from Jan 22 to Jan 30 was calculated, and the average incidence with the smallest coefficient variable was used to estimate the epidemic scale of COVID-19 in Wuhan during the same period.

**Figure 1.**
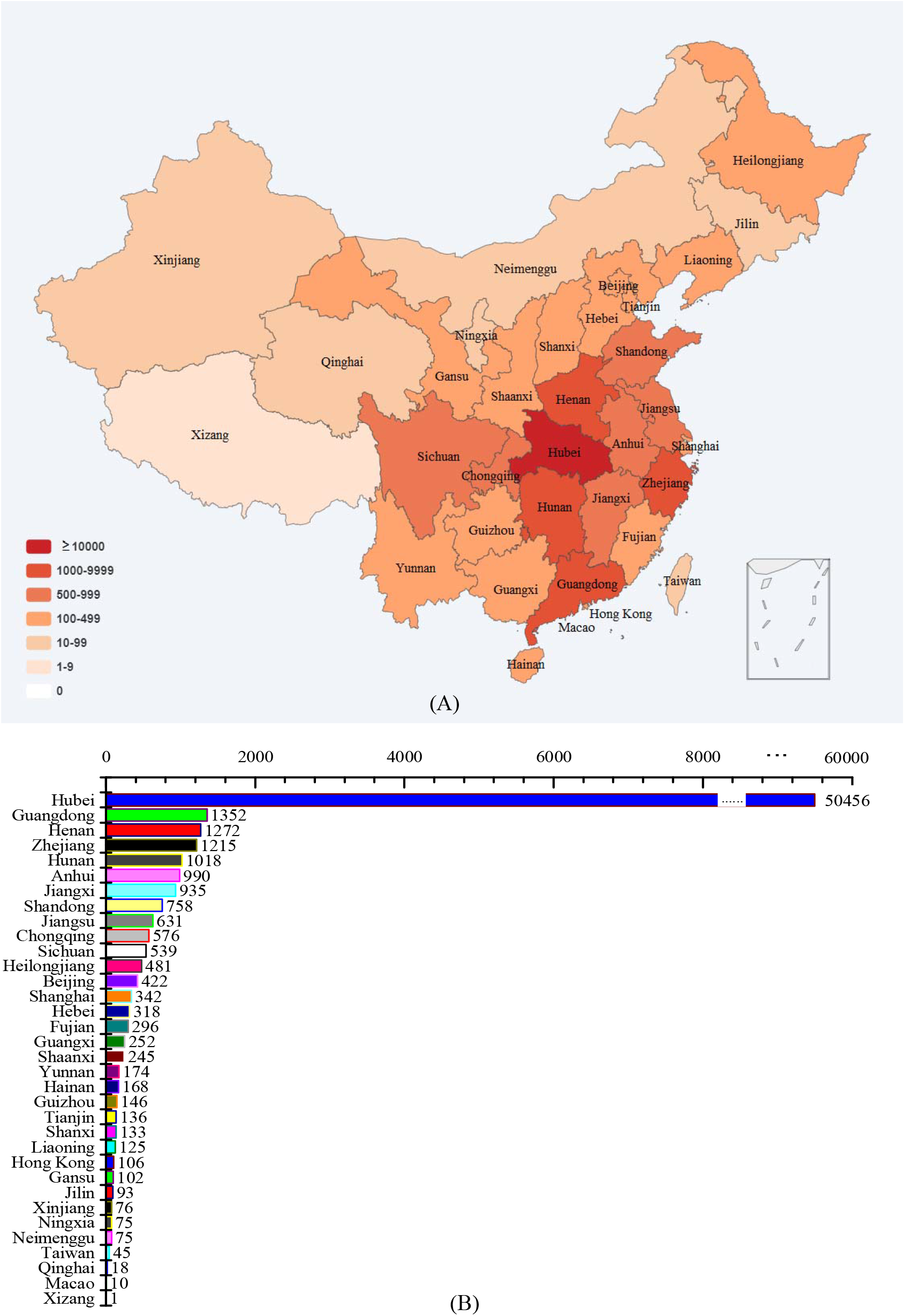
The epidemic distribution of COVID-19 in China (Mar 6, 2020). (A) the epidemic distribution map of COVID-19 in China; (B) the epidemic distribution of COVID-19 in the province of China.

### 2.2 Establishment of SEIRD model based on differential equations and estimation of basic reproductive number (R_0_)

the SEIRD model was constructed by adding the incubation period (E) and death (D) on the basis of the classical epidemic SIR transmission model [21-22]. The flow chart is presented as follows:

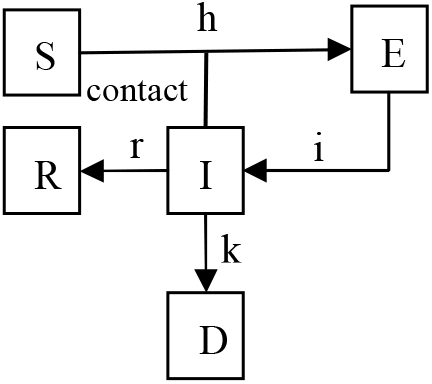

The variables are defined as follows: S: generally susceptible population; E: exposed population; T_E_: average incubation period; I: infectious population; T_I_: average duration of the infectious period; R: quarantined population (people who are cured and immunized or effectively quarantined); D: people who have died.

The used parameters are the following: h: the probability of a susceptible person (S) entering into the incubation period after coming in contact with an infectious person (I), h = R_0_/T_I_, where R_0_ is the basic reproductive number; i: the probability of a person in the incubation period (E) entering into the infectious period per unit time (I), which is the reciprocal of the incubation period, i = 1/T_E_; r: the probability of a person in the infectious period (I) entering into an isolated (R) state per unit time, which is the reciprocal of the duration of the infectious period or the course of disease, r = 1/T_I_; k: the probability of a person in the infectious period (I) to die per unit time, which is the product of mortality and recovery rate (r).

Therefore, the differential equations of transmission are as follows:

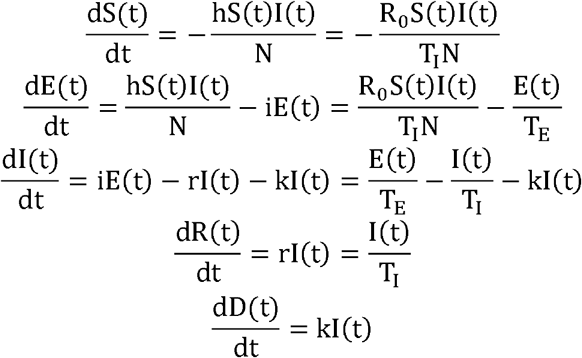

where S(t), E(t), I(t), R(t), and D(t) represent the number of susceptible, exposed, infectious, recovered, and dead individuals in the community at time t, respectively, and N represents the total number of individuals in the population, where N = S(t) + E(t) + I(t) + R(t) + D(t).

Because the population is very large, it can be assumed that S(t) = N, and the corresponding differential equations are as follows:

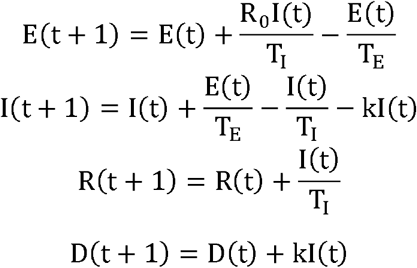

Therefore, the dynamic transmission process of SARS-COV-2 can be estimated by the upper differential equation.

R_0_ refers to the average number of secondary cases generated by one primary case in a susceptible population. According to the propagation dynamics [16], the formula is

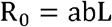

where a is the average daily contact time between an infectious patient and susceptible people, b is the probability of successful infection in each contact, and L is the time of transmission, which can be regarded as equivalent to T_I_.

### 2.3 The release time nodes of China’s public health interventions policies

The related forty public health interventions policies were obtained from the website of the National Health and Commission of China (Supplementary table 1).

**Table 1.** Comparison between the actual and simulated numbers of cases.

|  | Actual | Simulated |  |  | Actual | Simulated |  |
| --- | --- | --- | --- | --- | --- | --- | --- |
| Date | cumulative | cumulative | Relative | Date | cumulative | cumulative | Relative |
|  | cases | cases | error% |  | cases | cases | error% |
| Jan 30 | 9692 | 14773 | 52.4 | Feb 17 | 55053 | 54559 | 0.9 |
| Jan 31 | 11791 | 16825 | 42.7 | Feb 18 | 56855 | 55969 | 1.6 |
| Feb 1 | 14380 | 18992 | 32.1 | Feb 19 | 57675 | 57289 | 0.7 |
| Feb 2 | 17205 | 21293 | 23.8 | Feb 20 | 58564 | 58523 | 0.1 |
| Feb 3 | 20438 | 23745 | 16.2 | Feb 21 | 58961 | 59678 | 1.2 |
| Feb 4 | 24324 | 26359 | 8.4 | Feb 22 | 59609 | 60760 | 1.9 |
| Feb 5 | 28018 | 29149 | 4.0 | Feb 23 | 60018 | 61772 | 2.9 |
| Feb 6 | 31161 | 31778 | 2.0 | Feb 24 | 60526 | 62720 | 3.6 |
| Feb 7 | 34546 | 34294 | 0.7 | Feb 25 | 60932 | 63608 | 4.4 |
| Feb 8 | 37198 | 36724 | 1.3 | Feb 26 | 61365 | 64439 | 5.0 |
| Feb 9 | 40171 | 39083 | 2.7 | Feb 27 | 61692 | 65218 | 5.7 |
| Feb 10 | 42638 | 41382 | 2.9 | Feb 28 | 62119 | 65947 | 6.2 |
| Feb 11 | 44653 | 43625 | 2.3 | Feb 29 | 62692 | 66629 | 6.3 |
| Feb 12 | 46472 | 45815 | 1.4 | Mar 1 | 62894 | 67268 | 7.0 |
| Feb 13 | 48467 | 47831 | 1.3 | Mar 2 | 63019 | 67867 | 7.7 |
| Feb 14 | 49970 | 49698 | 0.5 | Mar 3 | 63138 | 68427 | 8.4 |
| Feb 15 | 51090 | 51433 | 0.7 | Mar 4 | 63277 | 68952 | 9.0 |
| Feb 16 | 53149 | 53049 | 0.2 | Mar 5 | 63420 | 69444 | 9.5 |

### 2.4 The model parameter assumptions and model fitting

The public health interventions are becoming increasingly strong. As of Jan 29, 31 provinces have launched a first-level response mechanism for public health emergencies. It can be assumed that Jan 30 is the node for strengthening public health interventions. From the policies of Feb 5, it can be assumed that it is another node at which public health interventions were strengthened. Therefore, we consider the period before the closure of Wuhan on Jan 23 as the pre-control stage, the period from Jan 24 to Jan 29 as the transition stage, and the period from Jan 30 as the post-control stage.

The model parameters are assumed as follows. (1) Due to the approaching Spring Festival, the number of contacts per unit time during the first two days of Wuhan’s closure (Jan 22 and Jan 23) increased by 200%. (2) Due to the publicity of the epidemic, the patients with mild symptoms obtained medical treatment in a timely manner since Jan 23, which reduced the duration of the infectious period from 4 days to 3.5 days and reduced L by 13%. And the number of contacts per unit time dropped by 20%. (3) On Jan 30, Measures such as village road closures and public segregation have been implemented throughout the country, reducing the number of contacts per unit time by 40%. (4) As a result of wearing masks, disinfection, and other preventative measures, the number of contacts per unit time was reduced by 60%, and the probability of single contact infection decreased by 30% since Feb 5. (5) After Feb 11, the number of clinically confirmed cases in Hubei Province was included in the number of confirmed cases by the National Health Commission of China, which would be more conducive to the control of the epidemic [23]. Therefore, three states (A, B, and C) were assumed: under state A, R dropped to 0.5 from Feb 12; under state B, R dropped to 0.9 from Feb 5 and to 0.5 from Feb 20; and under State C, R dropped to 0.9 from Feb 5 and to 0.5 from Mar 5.

### 2.5 Analysis of model effectiveness

In order to analyze the fitting degree of simulated data, the relative error of the actual cases and simulated cases was calculated. In addition, the relevant index (H^2^) was calculated by the following formula:

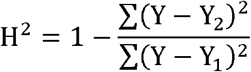

where Y is the number of actual cases, Y_1_ is the average of the number of actual cases, and Y_2_ is the simulated number of cases.

### 2.6 Evaluation of public health interventions of China on the transmission dynamics of SARS-COV-2

On the one hand, the reduction of reproductive number of the simulate curve was calculated. On the other hand, the infection curve, the daily transmission replication curve, and the cumulative confirmed cases trend curve were used to evaluate the impacts of public health interventions of China on the transmission dynamics of COVID-19 caused by SARS-COV-2. The infection curve was obtained by adding an incubation period to the curve of newly confirmed cases. A daily transmission replication number (R_t_) curve was constructed. The number of infections in n + 4 days (according to the infection period T_I_ = 4) is the number of people re-infected by the source of infection in n days, and the ratio was calculated. Because accelerating the speed of patient admission and shortening the time from symptom onset to isolation have a significant effect on epidemic control [23], the trend curve of cumulative confirmed cases was also used.

## 3. Results

### 3.1 Estimation of the basic reproductive number (R_0_) and the transmission curve fitting model

The average incidence of 34 cases (95% CI, 25.3–42.9) per 10,000 people moving out from Wuhan on Jan 28 was selected as a reference for estimating the incidence of COVID-19 in Wuhan. Based on this incidence, we estimated that the average number of COVID-19 cases during the incubation period in Wuhan from Jan 14 to Jan 21 was about 5,586 (95% CI, 4,156–7,048). The incubation period of COVID-19 is 2–14 days [12], and T_E_ is about 6 days [11]. The T_I_ (refer to the relevant studies of SARS [16, 24]) is 4 days, and the mortality is 0.0306, so the daily mortality is 0.0306/4 = 0.00765. According to the WMHC, the first hospitalized case of COVID-19 was recorded on Dec 8, 2019, and we estimate the first infected patient appeared on Dec 1, 2019. Therefore, the time series of E(t), I(t), R(t), and D(t) were calculated by four iterative equations, and the number of people in the incubation period in Wuhan from Jan 14 to Jan 21 is 5,586, and R_0_ is 3.38 (95% CI, 3.25–3.48).

Based on the assumed parameters of model, R was reduced to 2.37 on Jan 24 and to 1.78 on Jan 30 (because 3.38 × (1 − 0.40) × (1 − 0.13) = 1.78). On Feb 5, R decreased to 0.83 (because 3.38 × (1 − 0.60) × (1 − 0.30) × (1 − 0.13) = 0.83). The parameters of the fitting curves of the SEIRD model were as follows: from Jan 24 to Jan 29, R = 2.37, T_E_ = 6, T_I_ = 3.5, and k = 0.0306; from Jan 30 to Feb 4, R = 1.78, T_E_ = 6, T_I_ = 3.5, and k = 0.0306; and from Feb 5 to Feb 11, R = 0.83, T_E_ = 6, T_I_ = 3.5, and k = 0.0306.

States A, B, and C are described in the Methods section. State D is the natural transmission mode. State E is the transmission mode in which public health interventions are not strengthened after Jan 24. State F is the transmission mode in which public health mesures are not strengthened after Jan 30. All these state fitting curves are shown in Figure 2. It can be observed that the actual number of cumulative confirmed cases before Feb 12 remains below the simulated curve. For state A, it is expected that the number of cumulative cases of COVID-19 will reach about 74,000. State B is expected to reach about 89,000, and state C is expected to reach about 100,000.

**Figure 2.**
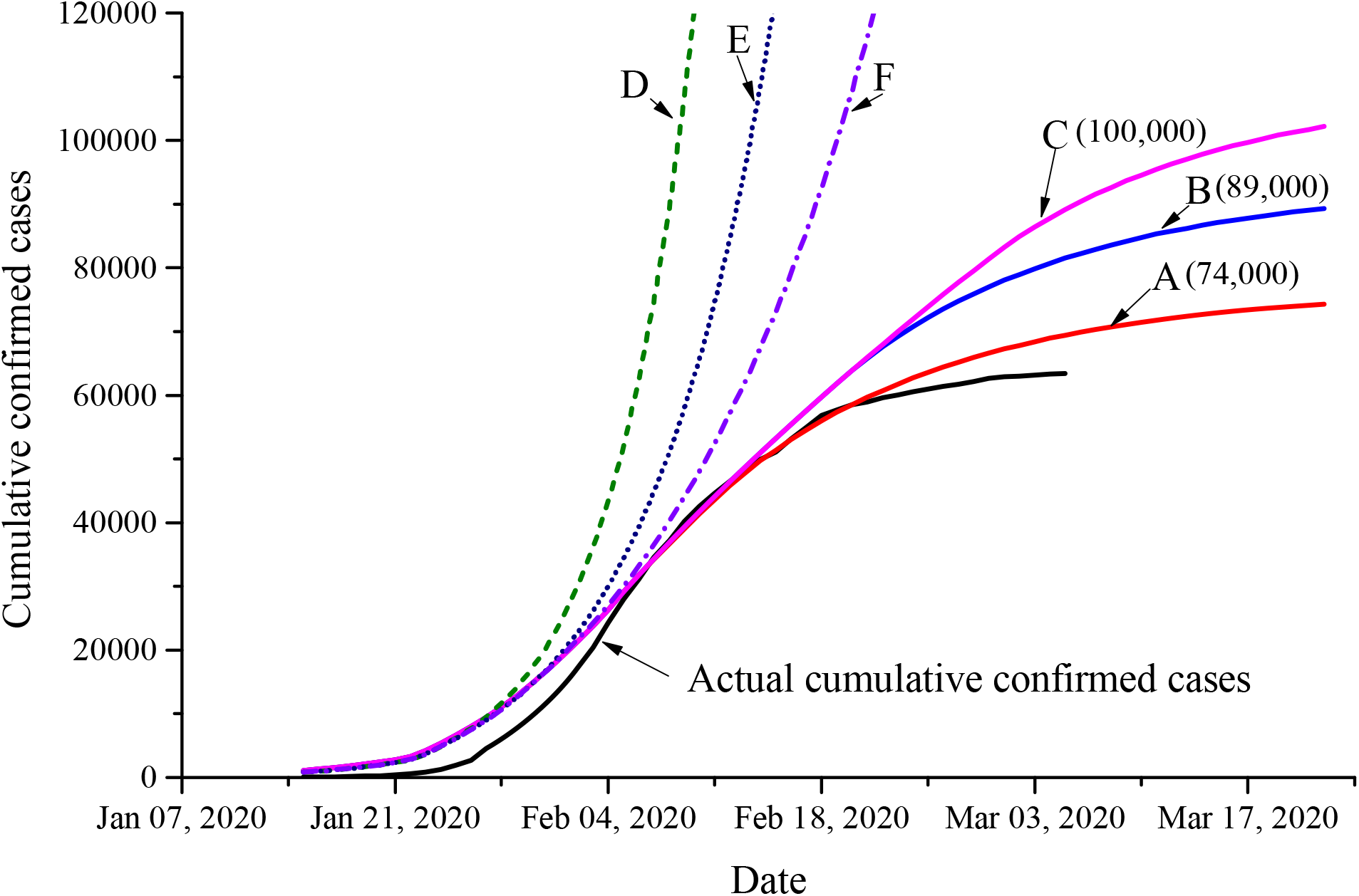
Simulation and prediction trend of the cumulative number of COVID-19 confirmed cases.

The difference between the simulated number of daily new cases and the actual number is shown in Figure 3. The number of daily actual new cases is lower than the simulated number before Feb 1 (except Jan 27), and the simulated number of daily new cases reached a peak of 2,790 on Feb 5. However, the actual number of daily new cases reached a peak of 3,886 on Feb 4.

**Figure 3.**
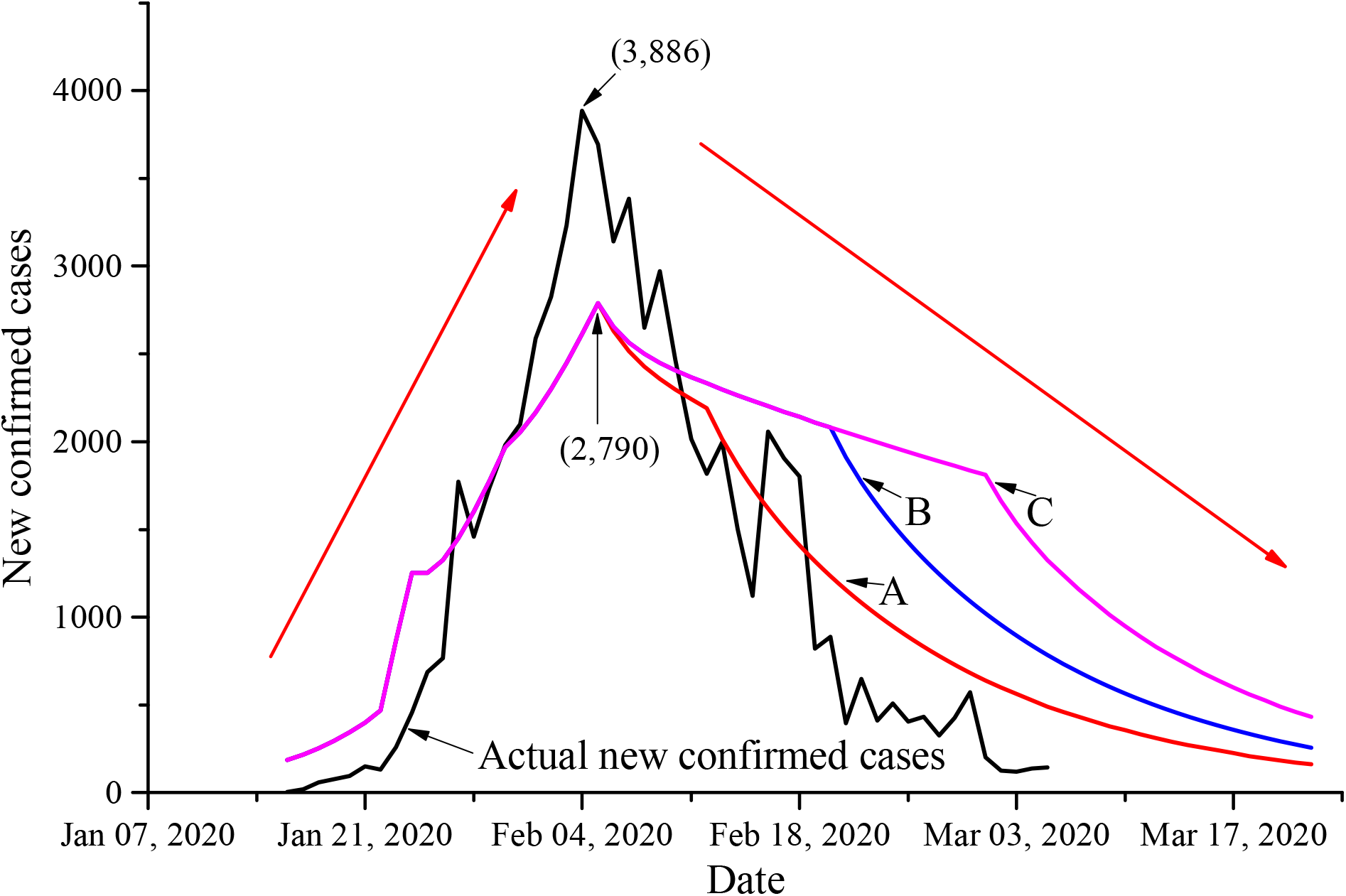
Simulation and prediction of the number of new COVID-19 confirmed cases.

The curves of the simulated cumulative number of deaths and the actual cumulative number of deaths in each state are shown in Figure 4. The actual number of new cases before Feb 10 is lower than the simulated number. For state A, it is expected that the number of cumulative deaths by COVID-19 will reach about 2,000. State B is expected to reach about 2,600, and state C is expected to reach about 3,000.

**Figure 4.**
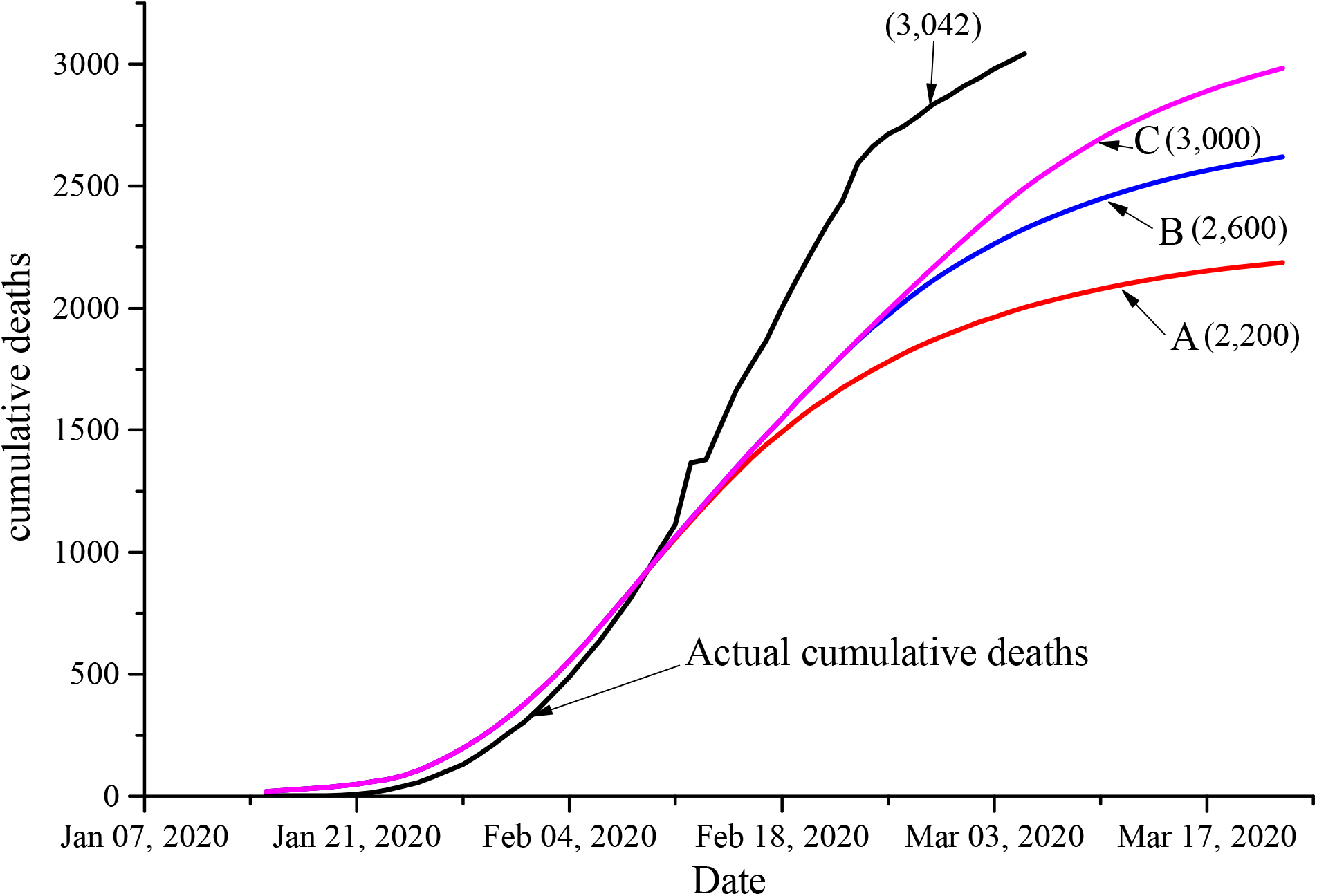
Simulation and prediction of cumulative deaths caused by COVID-19.

### 3.2 Analysis of model effectiveness

From Jan 30 to Feb 4, the maximum relative error is 52.4%, and the average relative error is 29.3%, as shown in Table 1. However, with the improvement of the diagnostic conditions (input from various temporary hospitals), the actual cumulative number of cases was closer to the simulated number from Feb 5 to Mar 5. The maximum relative error was 9.5%, and the average relative error was 3.4%. The H^2^ of the number of cumulative cases between Feb 5 and Mar 5 is 0.934.

### 3.3 Evaluation of public health interventions of China on the transmission dynamics

The incubation period (T_E_ = 6) was subtracted from the incidence curve to obtain the infection curve, and the actual data were added to the incubation period data according to the SEIRD model described above (Figure 5). The peak of infection occurred on Jan 29, and the outbreak has been controlled since that day. The number of daily infections has decreased significantly, which indicates that the current public health mesures of China are effective.

**Figure 5.**
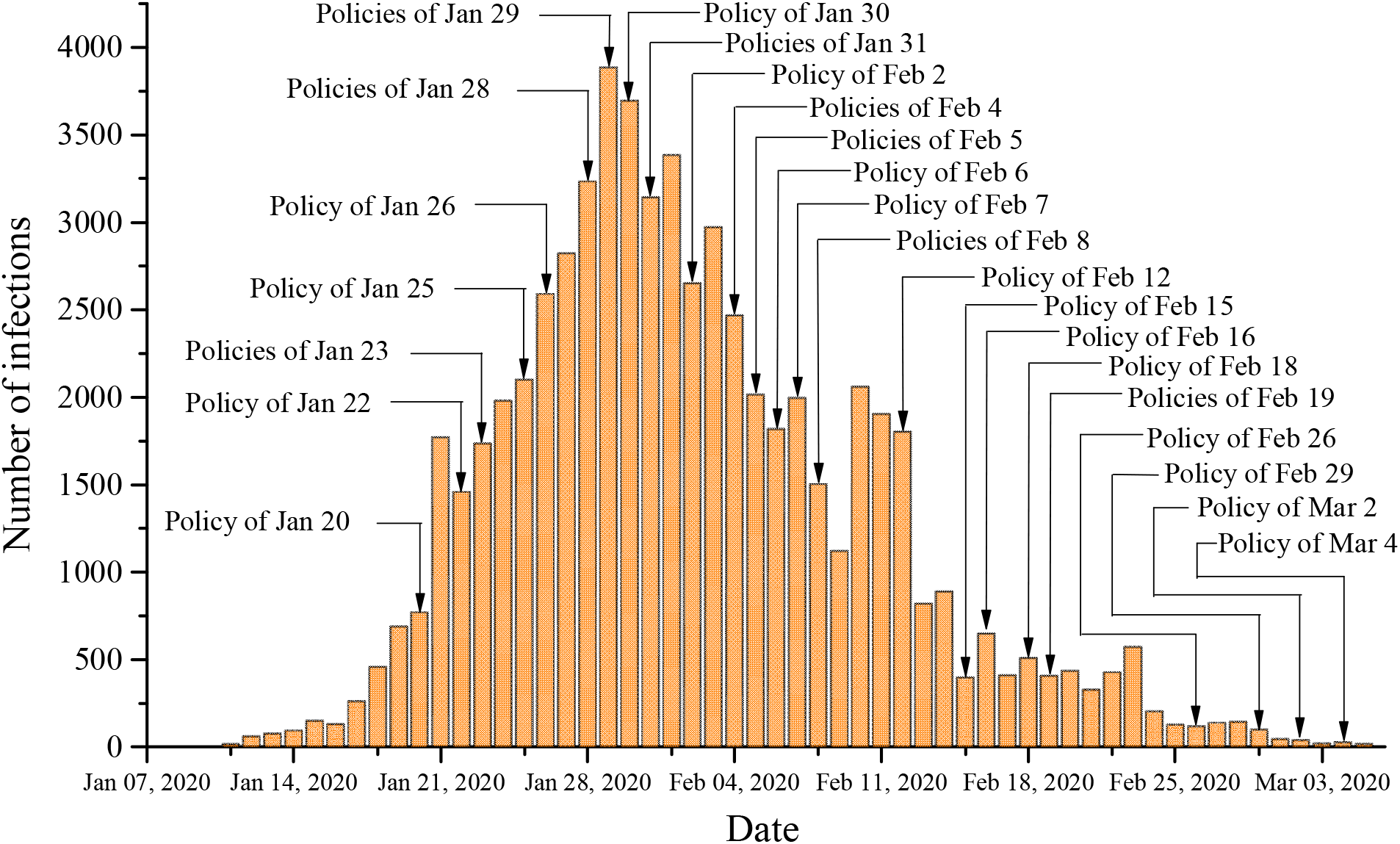
Daily infection situation of SARS-COV-2.

The transmission replication curve is shown in Figure 6. The rate of transmission and replication is very high and fluctuates greatly before Jan 23, with an average R of 3.20 (95% CI, 2.28–4.12), which may be due to the partial distortion of the number of previously reported cases. The average number of transmission and replication decreased to 1.5 between Jan 24 and Feb 5, which indicates that the epidemic was under control. It fluctuates around 1 since Feb 15, which may be related to the people returning to work in China. The rapid decline of the average re-infection rate indicates that multiple public health interventions have worked simultaneously, and the impacts are positive.

**Figure 6.**
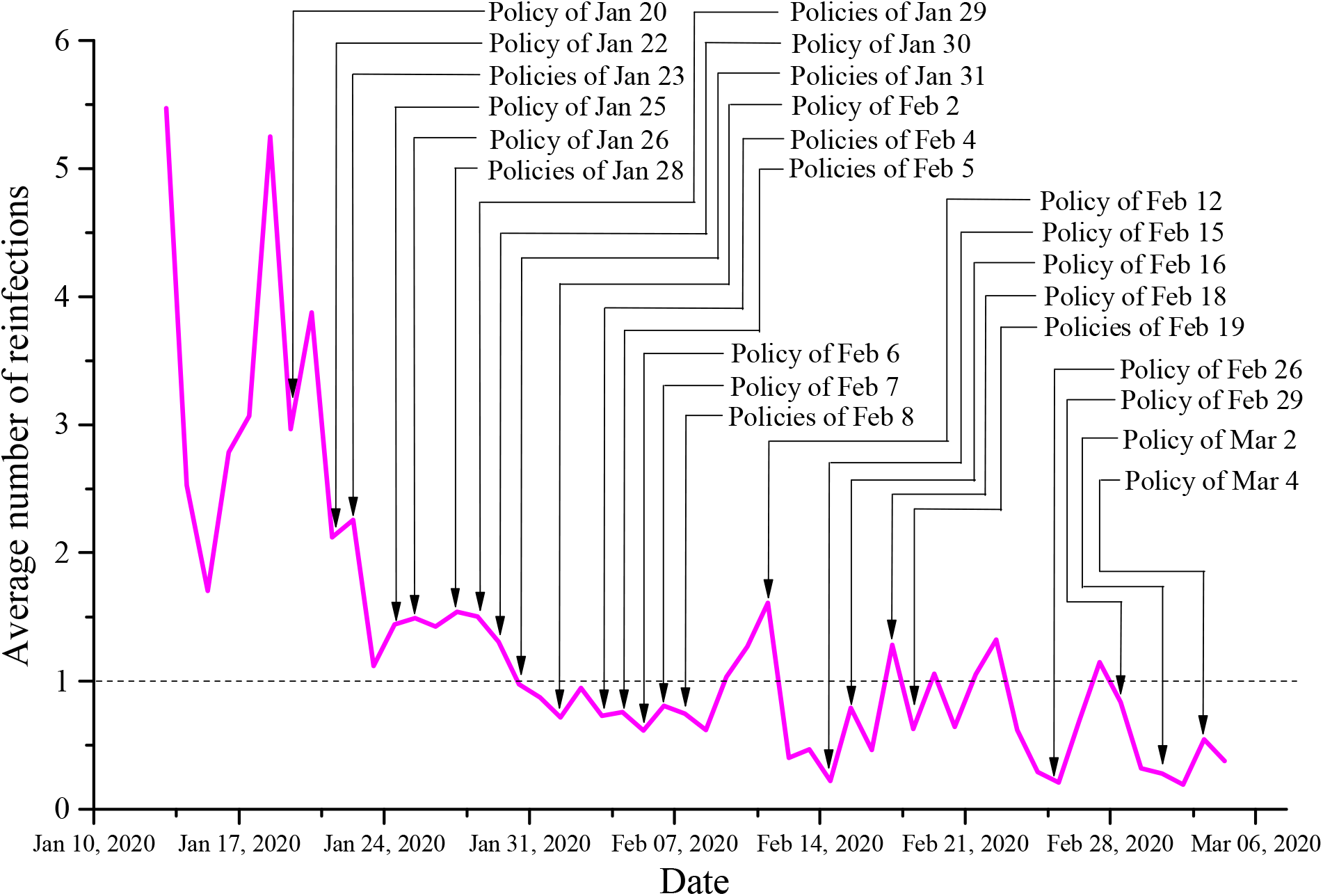
Curve of average transmission replication of SARS-COV-2.

The number of clinically confirmed cases in Hubei Province are included in the number of confirmed cases from Feb 12, which will be more conducive to the control of the epidemic and shorten the remaining time of the epidemic. The trend of cumulative confirmed cases after shortening the incubation period is shown in Figure 7. The final estimated cumulative confirmed cases (including clinical diagnosis) will be between curve A and curve B, and the epidemic will end soon in China.

**Figure 7.**
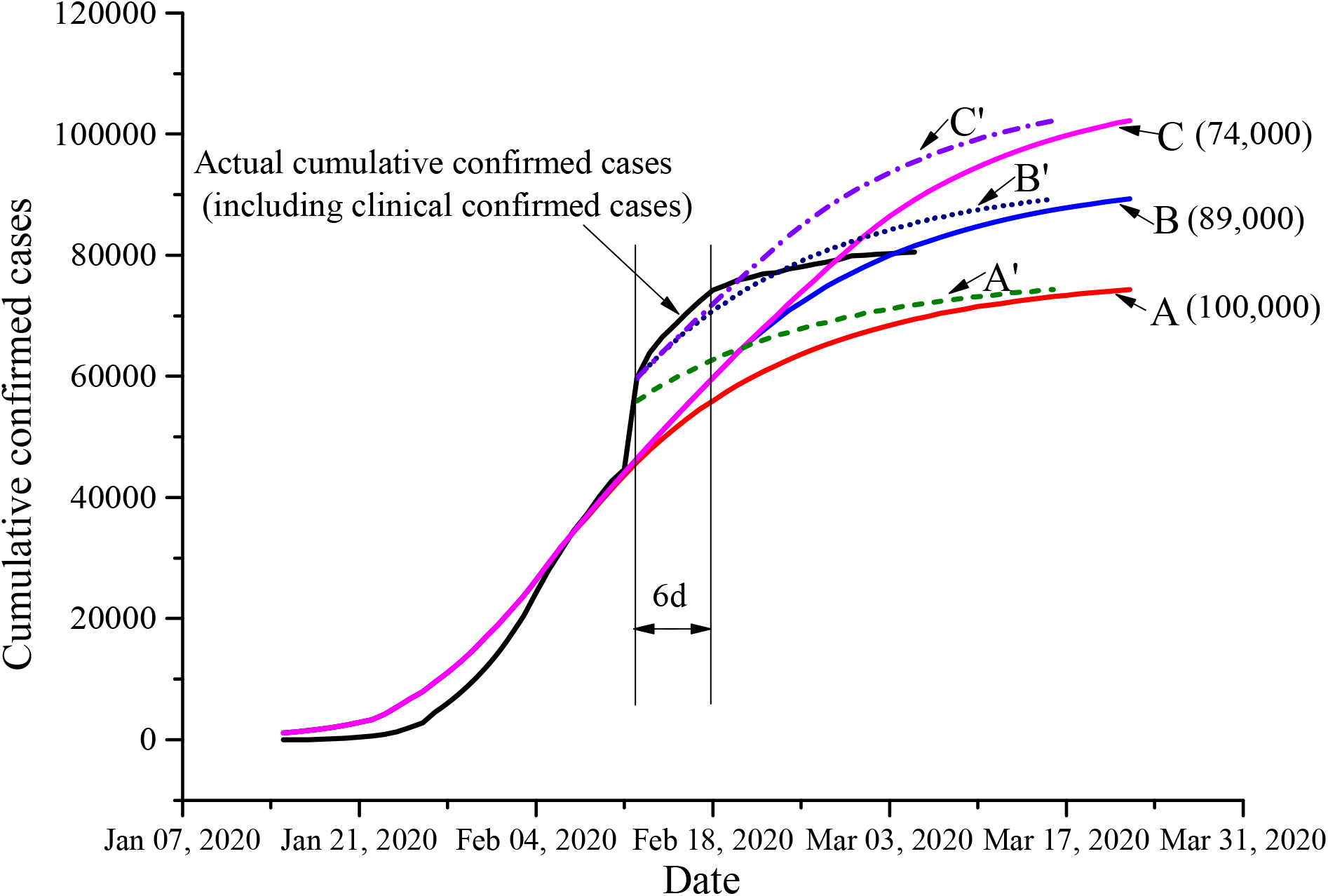
Trend of cumulative confirmed cases (including clinically confirmed cases).

## 4. Discussion

For a new infectious disease that can be transmitted from human to human, understanding its transmission dynamics is crucial for the formulation and prevention of public health interventions [25]. Excessive public health interventions have serious impacts on social and economic activities and people’s daily lives, while inadequate measures will cause epidemic spread and pose a huge threat to global health. The establishment of appropriate mathematical models to predict the spread of infectious diseases is particularly important for the prevention and control of infectious diseases [14-15]. SIR, SEIR, and SEIRD are commonly used mathematical models for infectious diseases [16-19]. Because missed diagnosis and misdiagnosis are inevitable in the emergence of a new infectious disease, the SEIRD five-compartment model is more credible than the others. Therefore, in the present study, a SEIRD propagation dynamics model was constructed to simulate the Wuhan epidemic since its emergence in Dec, 2019.

The SEIRD model contains three main parameters: R_0_, T_E_, and T_I_. Therefore, calculating R_0_ is crucial to estimate the transmission dynamics of COVID-19 caused by SARS-COV-2. For the determination of R_0_ of new infectious diseases, the simplest method is to estimate the multiplication time of the epidemic in the early stage, then to take into account the random volatility in the early stages, and finally to fit the exponential growth equation to calculate it. However, results obtained following this method may have limited accuracy because other factors are not taken into account [26]. There also lies a certain uncertainty in Wuhan’s early epidemiological data to estimate R_0_, although a previous study had estimated R_0_ is about 2.2 [12]. Therefore, in the present study, we used the incidence of COVID-19 among the population who moved out of Wuhan to eight provinces, and then reverse estimated the epidemic scale of COVID-19 in Wuhan during the same period. The estimated value for R_0_ (3.38, 95% CI, 3.25–3.48) is similar to that of SARS in Singapore and Hong Kong, which at the beginning ranged from 2 to 4 [16-17]. Previous studies estimated that R_0_ is between 2.2 and 2.68 [11-12], which is lower than the results of the present study. This may be related to the limited early epidemical data and the lagging diagnosis in the early stage in Wuhan. However, after Jan 22, the diagnosis methods became more reliable, and coupled with the closure of Wuhan and the government’s attention, the public’s awareness of epidemic prevention improved, resulting in a relatively accurate measurement of COVID-19 incidence. Therefore, we consider that the value for R_0_ of the present study may be more accurate.

In the SEIRD model we constructed, if R is not controlled, the number of cases will increase exponentially, as shown in curve D in Figure 3. Quarantine measures can reduce the possibility of contact with infected individuals before the onset of disease symptoms [16, 27], and real-time monitoring of quarantined individuals can shorten the time from symptom onset to hospital admission. In addition, changing the behavior of the public, such as reducing the frequency of visits to public places and reducing the use of public transportation, can reduce the chance of contact and thus the value of R [17]. On Jan 23, the Chinese government carried out public health interventions (closing the city, reducing the frequency of visits to public places, reducing the use of public transport, and other measures) to make R less than R_0_. At this time, the cumulative number of cases changes from curve D to curve E. With the strengthening of the public health interventions and the deepening of the propaganda of the epidemic, the public took personal protective measures, such as wearing masks and gloves and washing hands frequently, to reduce the effective infection rate [23, 27-28]. After Jan 30, the cumulative curve of cases is reduced from curve E to curve F. Another method to reduce R is to shorten the average infection duration of the disease, which comes down to reducing the natural infection duration of the host through the symptomatic treatment with antiviral agents or reducing the effective duration of infection by early detection, early isolation, and early treatment of the disease. Based on the SARS epidemic, many studies have pointed out that shortening the time from the onset of symptoms to the isolation of cases has a significant effect on the control of the large-scale epidemic of the disease [16-17, 23]. With the introduction of related policies on quarantine measures, R was below 1 from Feb 5. This indicates that not each case of infection infects a second-generation case, and the epidemic will end in the near future.

Our fitting model showed that from Jan 24 to Jan 29, the actual cumulative number of cases was much lower than the simulated cumulative number (Figure 3), and the actual number of new cases was much lower than the simulated number (Figure 4), which may be related to the lack of conditions for diagnosis or the low efficiency of diagnosis in some early grassroots hospitals in Wuhan, as well as the late use of diagnostic test kits. From Jan 30 to Feb 4, with the increased supply of test kits, the gap between the actual cumulative number of cases and the simulated cumulative number of cases became smaller. Because the H^2^ value was closer to 1, the actual number of cases was closer to the simulated number, and the curve fitting was better. The low relative error and the high H^2^ value of 0.934 in the present study illustrate the accuracy and reliability of the SEIRD model. At present, although methods to quantitatively evaluate public health interventions are lacking, the combination of daily infections and daily transmitted replication with interventions can directly reflect the effects of intervention policies on daily infections or daily transmitted replication and accurately reflect the effectiveness of intervention policies. In the present study, time series analysis combined with an incubation period was used to draw the infection curve. At the same time, the daily replication curve was used to visualize the change in daily infection status, analyze dynamic infection information, and evaluate the effects of control measures. The time series analysis curve, the daily transmission replication curve, and the confirmed cases trend chart reflect the change of infection status from different perspectives. The results of the three methods are consistent, suggesting that the intervention measures are effective.

There are some limitations in this study. For example, the source of infection of COVID-19 is not completely clear, and the model assumes that the transmission of COVID-19 only takes place from human to human; the model assumes that the population is uniformly mixed, and that each individual in the same population has the same daily contact rate with the patients and the same probability of infection upon one close contact. In addition, the model does not take into account the climate, especially the possible effects of temperature on the transmission of COVID-19. Despite these limitations, our modeling results are in good agreement with the actual numbers, and to a certain extent, the effects of public health interventions are well evaluated. Our study provides a theoretical framework for the formulation and implementation of follow-up public health prevention and control measures. At the same time, it can also provide a reference for the world to deal with the current pandemic.

## 5. Conclusions

Our results suggest that the transmission dynamics of SARS-COV-2 of China were now decline and the current public health interventions are effective and should be maintained until COVID-19 is no longer considered a global threat.

## Data Availability

All data referred to in the manuscript is available by the corresponding author.

## Supplementary Materials

Table S1: The public health intervention policies of China (Jan 20 until Mar 4, 2020)

## Author Contributions

W.bW. contributed to the study design, model establishment, data analysis, and the write-up. Y.C. and Q.W. contributed to the figure and table creation. P.C. and Y.H. contributed to collect additional required data specific to China, and provide policy-relevant insight. S.H. and Y.W. contributed to the data analysis and data interpretation. W.xW. oversaw all aspects of study design, model evaluation, data analysis, data interpretation, figure design, and the write-up.

## Acknowledgments

We thank LetPub (www.letpub.com) for its linguistic assistance during the preparation of this manuscript.

